# Enhanced Demographically Adaptive QT Correction Improves Pediatric Screening for Congenital Long QT Syndrome

**DOI:** 10.64898/2026.05.14.26353243

**Authors:** Kazi T. Haq, Charles I. Berul, Nikki Gillum Posnack

## Abstract

**Background:** Traditional heart rate (HR) adjusted QT correction (QTc) formulae often fail to eliminate the inverse HR-QT interval relationship, particularly in pediatric patients. In this study, we optimized our previously published adaptive QTc (QTcAd) formula by including additional demographic variables and broadening the pediatric age range. We tested the hypothesis that QTcAd improves congenital long QT syndrome (congenital LQTS) detection performance and reduces erroneous classifications across pediatric cohorts.

**Methods:** We retrospectively analyzed 8,306 ECGs from 4,556 cardiovascular disease (CVD)-free pediatric patients. For neonatal patients (1-30 days old), we derived daily QTcAd parameter values. For older patients, we developed regression models to estimate QTcAd parameters (mean Heart Rate (HR) =-15.9ln(days) + 219; |m| = 0.0001(days) + 1, where |m|=absolute HR-QT regression slope). To support LQTS screening, we constructed dynamic QTcAd thresholds by estimating age-specific reference limits. Diagnostic performance was tested in a clinically confirmed LQTS cohort (n=137), and further evaluated in the Pediatric Heart Network (PHN; n=2,394) and Emergency Department (ED; n=2,002) cohorts.

**Results:** Using the confirmed LQTS cohort as the event population and the CVD-free cohort as the non-event population, QTcAd demonstrated higher sensitivity than QTcB (92% vs 46.7%). QTcAd maintained high specificity (96.9% vs 98.9%), which resulted in a higher Youden index (0.889 vs 0.456). In the PHN healthy cohort, both QTc formulae classified the majority of individuals as normal (QTcAd 95%; QTcB 98.2%) indicating few false-positives. In the ED cohort, QTcAd reduced borderline/prolonged QTc classifications requiring follow-up, yielding 270 fewer repeat-testing triggers than QTcB. We developed a publicly accessible calculator to compute QTcAd and classify congenital LQTS risk.

**Conclusion:** We developed and validated an enhanced QTcAd formula for pediatric patients. QTcAd-based-age-adjusted dynamic thresholding improved performance for congenital LQTS screening, while maintaining high specificity. This reduces false-positive LQTS classifications and repeat ECGs, thereby decreasing unnecessary downstream clinical evaluation.

**Graphical Abstract:** 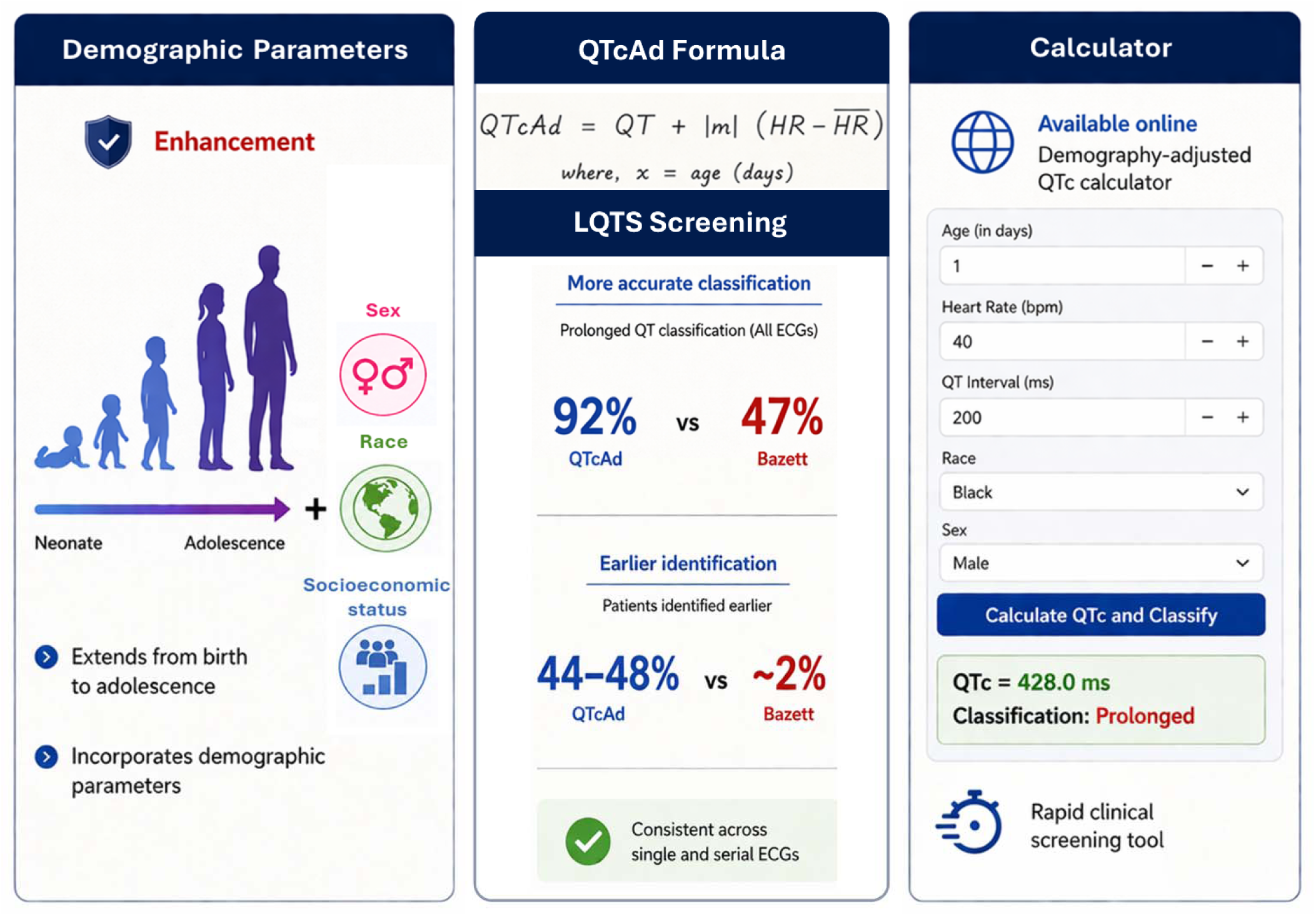

## Introduction

The heart rate-corrected QT interval (QTc) is an important electrocardiogram (ECG) biomarker that is widely used in cardiology. QTc prolongation above clinically defined thresholds is linked to serious cardiac risks, including congenital long QT syndrome (LQTS) and drug-induced delays in cardiac repolarization. Without treatment, these conditions can result in life-threatening events such as cardiac arrest or sudden cardiac death. QTc is frequently monitored to assess QT prolongation resulting from acquired cardiac disorders, electrolyte disturbances, or medications (i.e., antiarrhythmics, antibiotics, psychotropic drugs).

In the pediatric population, a QTc interval ≥460-480 ms is considered abnormal and may be attributable to congenital LQTS. The prevalence of congenital LQTS is estimated to be 1:2000 to 1:2500, but the incidence may be even higher due to variable penetrance and silent gene carriers (Schwartz et al., 2009; Vink et al., 2018). Despite the serious clinical implications of congenital LQTS, ECG-based screening methodologies can yield both false-positives and false-negatives in children (Andršová et al., 2020; Rodday et al., 2012). As an example, nearly 1/3 of children presenting at the emergency department with QT prolongation exhibited a normalized value in follow-up evaluations (Van Dorn et al., 2011). One factor that may be contributing to LQTS misdiagnosis is a lack of age-adjustment in QTc thresholds or the modified Schwartz score (Johnson & Ackerman, 2009; Leppänen et al., 2025; Schwartz & Crotti, 2011). Since QT and QTc intervals are strongly influenced by patient age (Benatar & Feenstra, 2015; Haq et al., 2023, 2025; Pærregaard et al., 2021), an age-adjusted QTc formula may improve the accuracy of congenital LQTS screening.

To date, several QT correction formulae (e.g., Bazett, Fridericia, Framingham, Hodges (Bazett, 1920; Fridericia LS, 1920; Hodges MS, 1983; Sagie et al., 1992)) are widely used to adjust the QT interval for heart rate. Nevertheless, QT correction formulae are often inaccurate across demographically diverse populations (Andršová et al., 2020; Benatar & Feenstra, 2015; Kwag et al., 2024; Malik, 2001; Malik et al., 2002). To address this limitation, we recently developed an adaptive QT correction formula (QTcAd) designed to incorporate age as a demographic variable and improve accuracy across diverse populations (Haq et al., 2025). However, the predictive efficacy of QTcAd in populations with cardiovascular conditions remains unexplored. In the current study, we incorporated additional demographic parameters (e.g., sex, race, socioeconomic status) alongside age and evaluated the performance of the enhanced QTcAd formula for congenital LQTS screening. To test whether the enhanced QTcAd formula would outperform Bazett QT correction (QTcB), we utilized retrospective ECG data collected from Children’s National Hospital (CNH) and the Pediatric Heart Network (PHN) (Saarel et al., 2018).

## Methods

### Study population

This study was conducted in accordance with the Declaration of Helsinki. This retrospective study (IRB #12146) included four pediatric cohorts **(Figure 1)**. (1) CNH CVD-free control cohort: A total of 4,556 patients contributed 8,306 ECGs. Patients were stratified based on ECG availability into single-ECG (n=3,064) and multiple-ECG (n=1,492) groups. The CNH CVD-free cohort included individuals aged 1 day-23 years, with equal distribution between males (50%) and females (50%), and the self-reported racial distribution included black (44%), white (26%), or all other races (30%). Socioeconomic characterization was evaluated using the Childhood Opportunity Index (COI 3.0) developed by Diversity Data Kids (https://www.diversitydatakids.org/child-opportunity-index) with a focus on the District of Columbia–Maryland–Virginia area, where most CNH centers are located. ZIP codes were collected for each patient and crossmatched to the COI 3.0 class using the state-normed method, with neighborhoods divided into five categories: very high (22%), high (19%), moderate (29%), low (9%), and very low (21%; **Figure 2A**). (2) CNH Clinically confirmed congenital LQTS cohort: A total of 410 patients with electronic health record-documented congenital LQTS confirmation were included. Across these patients, 3,140 ECGs obtained prior to diagnosis were available. The cohort was randomly divided at the patient level into a derivation subset (n=273 patients, n=1,991 ECGs) and an independent validation subset (n=137 patients, n=1,149 ECGs). (3) CNH Emergency department (ED) cohort: A total of 2,002 patients with a single ECG obtained during ED evaluation were included, reflecting a real-world screening environment. (4) PHN healthy cohort: This dataset included 2,394 individuals, with one ECG per patient.

**Figure 1.**
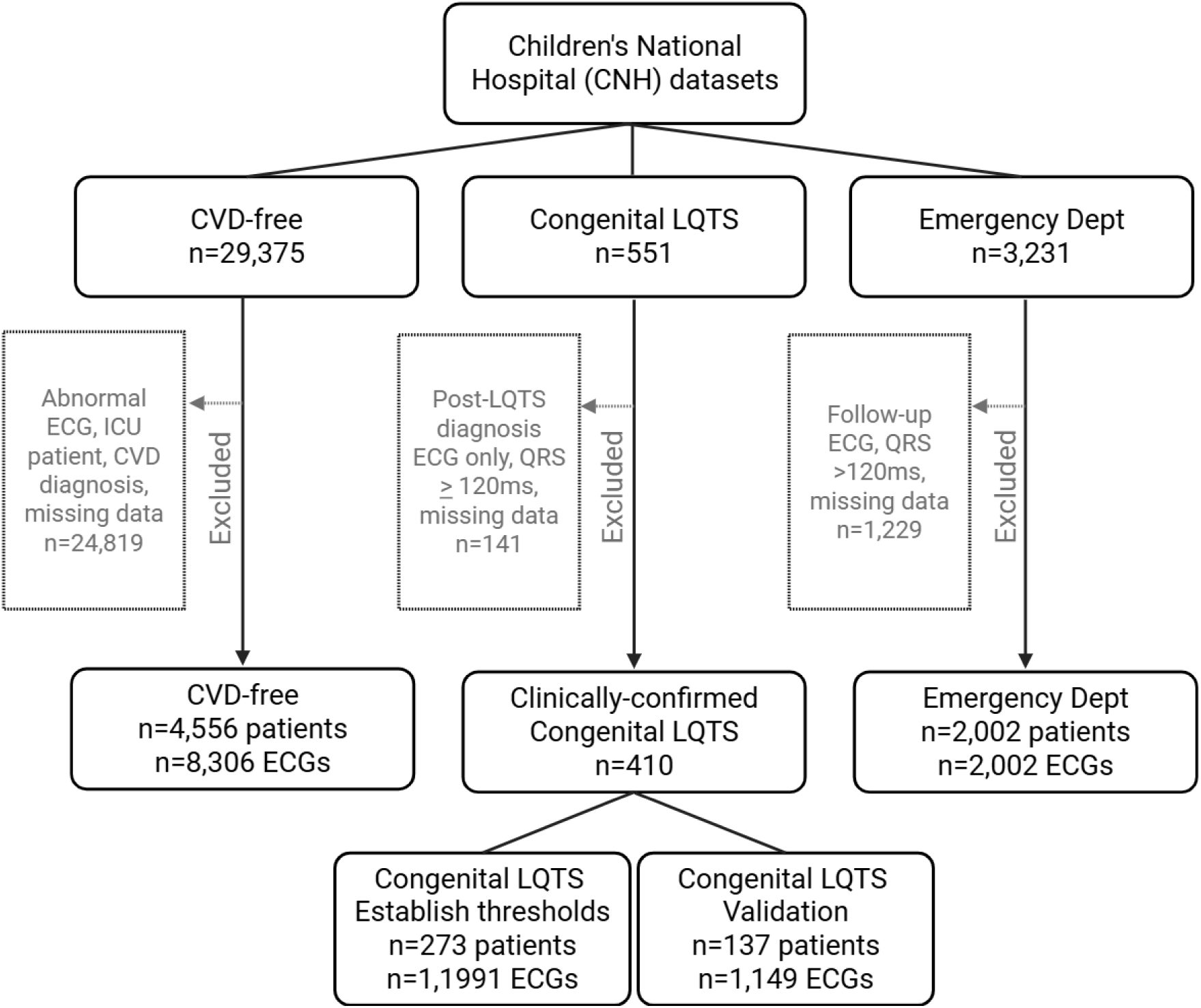
Children’s National Hospital cohorts. CVD-free cohort: Only included patients without a history of cardiovascular disease. Patients were also excluded if they were in an intensive care unit, data was missing, or their ECG was classified as abnormal. Congenital LQTS cohort: Only included patients with clinical confirmation of congenital LQTS. Patients were excluded if only a post-LQTS diagnosed ECG was available, data was missing, or the ECG showed an abnormal QRS interval. LQTS patients were subdivided into two cohorts to derive/establish LQTS threshold and to validate/test thresholds for correct diagnosis. Emergency Dept cohort: Only included patients visiting the emergency department. Follow-up ECG recordings were excluded.

**Figure 2.**
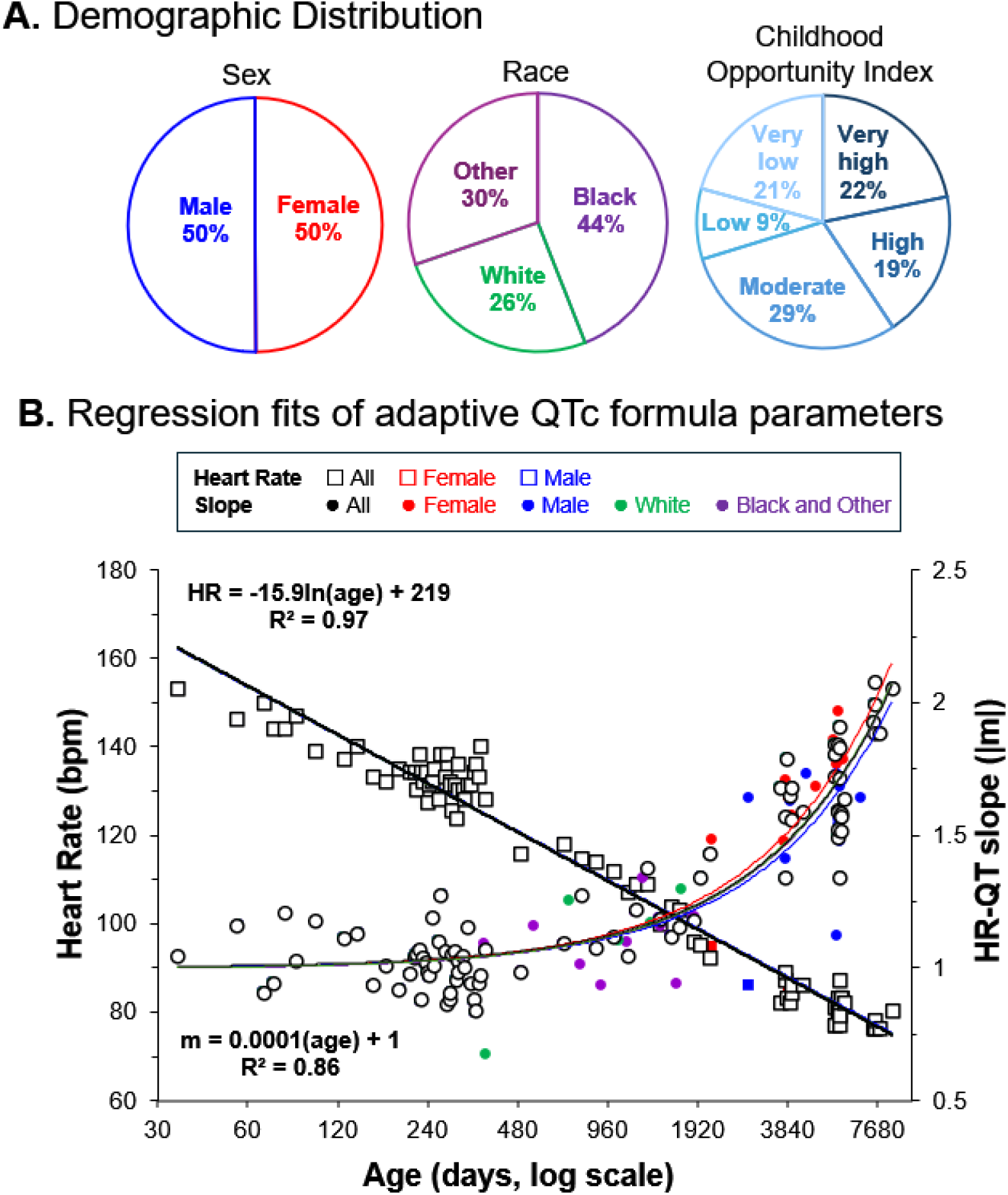
Patient demographic distribution and regression fits for the QTc formula parameters. **A.** Distribution of demographic characteristics in the CNH CVD-free cohort (n=4,556 patients). Sex categorized as male and female. Race categorized as white, black, or other, Childhood opportunity index categorized as very high, high, moderate, low, or very low. Percentages reflect the number of individual patients. **B)** Regression models predict heart rate and absolute value of heart rate-QT slope (m) in patients 1 month to 23 years old. Heart rate modeled by logarithmic fit for all CVD-free patients, males only (6-11 years), and females only (6-11 years). Slope modeled by linear regression fit for all CVD-free patients, males only (6-18 years), females only (6-18 years), or subdivided by race (white vs black/other, 1-5 years). Note: for clarity, only values that are statistically significantly different from the all CVD-free patient values are highlighted using different colors for female (red), male (blue), white (green), or black/other (purple). Age is displayed in days on a logarithmic scale. Regression equations and R² values are displayed within the plots.

## Exclusion criteria

### CNH datasets

*<u>CVD-free patients</u>* had no history of CVD (e.g., coronary artery disease, heart failure, arrhythmia, congenital heart disease etc.), pre/syncope, palpitations, or acute systemic illness (e.g. electrolyte disturbances, febrile seizures) at the time of ECG acquisition. Patients were excluded if the ECG report was classified as “abnormal” by a cardiologist. *<u>Clinically confirmed</u> <u>congenital LQTS patients</u>* did not have structural heart disease, electrolyte imbalance, medication use, or acute illness (e.g., fever, infection) at the time of ECG acquisition. *<u>ED</u> <u>patients</u>* did not have a history of arrhythmia, known or suspected congenital LQTS, CVD, or congenital heart disease, and were not taking medications known to prolong the QT interval at the time of ECG acquisition. For all patients, ECG datasets were excluded if there was significant noise, missing leads, signal quality issues, or physiologically implausible heart rates. ECGs with QRS ≥ 120 ms were excluded from any cohort included in congenital LQTS screening analysis.

### PHN dataset

This dataset is limited to health children, as detailed in (Saarel et al., 2018). This dataset excluded children with acquired or congenital heart disease, prematurity (<37 weeks gestation), children with obesity or an acute/systemic illness that could affect the cardiovascular system. This dataset also excluded children with a first-degree relative with nonischemic cardiomyopathy or obstructive congenital heart disease. ECGs were excluded if they did not meet digital waveform quality standards.

### Retrospective data collection

To build the CNH CVD-free cohort, we used the MUSE™ v9 ECG system and the Cerner PowerChart electronic health record database to collect data from 29,375 patients who visited a CNH center and underwent ECG testing between 1997 and 2024. The following data were collected: QT interval, HR, race, sex, age, residential zip code, ECG acquisition date, testing location, and a cardiologist’s interpretation of the ECG. Patients were only included if the cardiologist’s notes indicated a “normal” ECG waveform. Patients were excluded if they were admitted to an intensive care unit or had any history of cardiovascular disease. The final CNH CVD-free (control) cohort included 4,556 individuals aged 1 day–23 years. The CNH confirmed LQTS cohort included 410 patients, and the ED cohort included 2,002 patients. Patients were identified and their medical history was reviewed using the CNH database via Databricks (Databricks Inc.), an artificial intelligence-based data-mining platform [output reviewed by KH].

### Demographically enhanced QTcAd formula

We originally developed the QTcAd formula as (Haq et al., 2025) as:

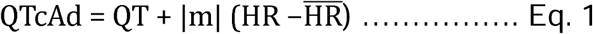

where |m| is the absolute value of the slope of the linear regression of the QT interval and HR, and 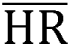 is the mean resting HR of a population.

Previously we utilized data from a pediatric population (1 day – 5 years old) to form a regression model to predict |m| and 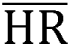 at a given age, using the following:

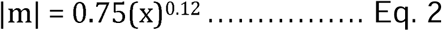

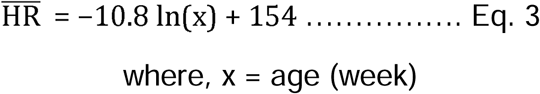

To enhance the QTcAd formula, we increased the age range from 1 day-5 years to 1 day-23 years. We also assessed whether other important demographic parameters (sex, age, childhood opportunity index) influenced the QTcAd formula expression and made additional adjustments accordingly in the final derivation.

In the CNH CVD-free cohort, we first assessed the effect of demographic parameters on the heart rate and QT interval using least square regression (**Table 1**). Heart rate and the QT interval were independently affected by age (p<0.001); but, sex, race, and childhood opportunity index were not independent predictors of either heart rate or the QT interval. However, a few of these demographic parameters interacted with age and/or other parameters to influence the ECG values. For example, in the neonatal age group, heart rate was significantly influenced by race (p<0.001; **Table 2**).

**Table 1.**
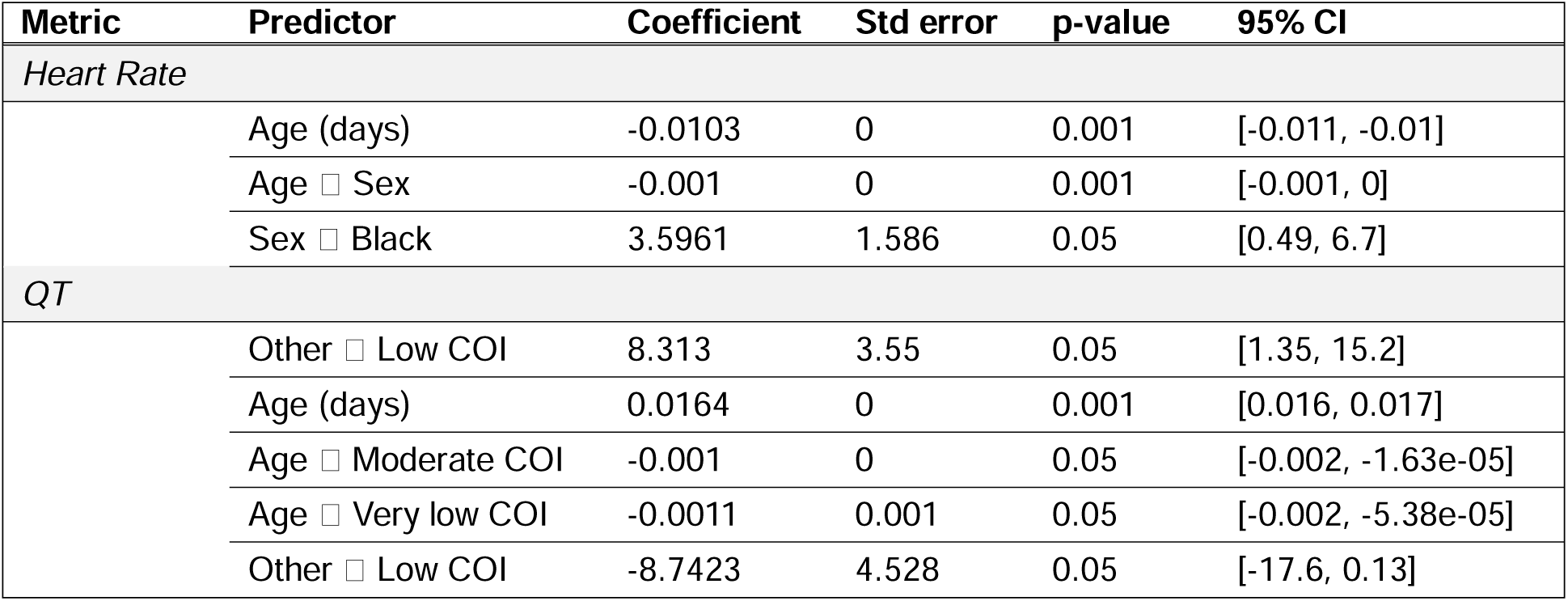
Effect of age, sex, race, and childhood opportunity index on heart rate and QT interval in the CNH CVD-free cohort. Patient age ranges from 1 day to 23 years. Sex categories include male or female. Race categories include white, black, and all other. Childhood opportunity index categories include very high, high, moderate, low, and very low. Statistical significance (ordinary least squares regression) is denoted by the p-value. CI: confidence Interval, Std error: Standard error

**Table 2.** Effect of sex, race, and childhood opportunity index on heart rate and QT intervals across six age groups. Patients were categorized into six age groups, including neonates (1-30 days), infants (31 days-12 months), early childhood (1-5 years), late childhood (6-11 years), early adolescents (12-18 years), late adolescents (19-23 years). Sex categories include male or female. Race categories include white, black, and all other. Childhood opportunity index categories include very high, high, moderate, low, and very low. Heart rate and QT interval were treated as continuous outcomes and analyzed using one-way ANOVA; statistical significance is denoted by the p-value.

| Metric | Age group | Predictor | F-value | p-value |
| --- | --- | --- | --- | --- |
| *Heart Rate* |  |  |  |  |
|  | Neonate | White □ Black | 12.47 | 0.001 |
|  |  | White □ Other | 5.96 | 0.05 |
|  | Infant | None | ns | ns |
|  | Early childhood | None | ns | ns |
|  | Late childhood | Female □ Male | 5.73 | 0.05 |
|  | Early adolescents | White □ Other | 4.21 | 0.05 |
|  | Late adolescents | High □ Low COI | 4.39 | 0.05 |
| *QT* |  |  |  |  |
|  | Neonate | White □ Black | 9.56 | 0.01 |
|  | Infant | None | ns | ns |
|  | Early childhood | White □ Other | 6.27 | 0.01 |
|  | Late childhood | None | ns | ns |
|  | Early adolescents | Female □ Male | 6.99 | 0.01 |
|  | Late adolescents | High □ Low COI | 6.79 | 0.01 |
|  |  | Low □ Very low COI | 5.29 | 0.05 |

Since neonates exhibit significant day-to-day changes in ECG metrics (Saarel et al., 2018), we calculated the |m| and 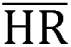 for each day from 1-30 days old (**Table 3**). Within each age, outlier ECGs were iteratively removed until the HR-QT interval correlation coefficient reached R^2^ > 0.6 to ensure a reliable fit; the minimum number of ECGs included per age (in days) was n=20.

**Table 3.** Adaptive QTc formula parameter values for neonates. . Heart rate (HR) and the absolute value of the HR-QT slope (|m|), two parameters of the QTcAd formula, were calculated for each day during neonatal life. Age values are presented in days. HR values are presented as bpm ± standard deviation. R² values represent the correlation coefficients from the linear regression of HR and QT. Number of samples is indicated by “n”. Interp: value obtained by linear interpolation between adjacent age-day estimates due to insufficient data (n≤5) for direct calculation in the specific age group.

|  | All patients |  |  |  | White |  |  |  | Black and Other |  |  |  |
| --- | --- | --- | --- | --- | --- | --- | --- | --- | --- | --- | --- | --- |
| Age | HR | $\vert m\vert $ | $R^2$ | n | HR | $\vert m\vert $ | $R^2$ | n | HR | $\vert m\vert $ | $R^2$ | n |
| 1 | 124 ± 27 | 1.24 | 0.85 | 22 | 129 ± 29 | 1.20 | 0.91 | 11 | 120 ± 25 | 1.29 | 0.78 | 11 |
| 2 | 132 ± 17 | 1.04 | 0.65 | 44 | 128 ± 18 | 1.00 | 0.61 | 20 | 135 ± 15 | 1.04 | 0.66 | 24 |
| 3 | 130 ± 15 | 1.08 | 0.70 | 33 | 130 ± 9 | 1.07 | 0.46 | 11 | 130 ± 18 | 0.95 | 0.64 | 24 |
| 4 | 127 ± 17 | 0.96 | 0.65 | 40 | 127 ± 10 | 1.58 | 0.74 | 15 | 126 ± 20 | 0.88 | 0.68 | 25 |
| 5 | 132 ± 22 | 1.00 | 0.85 | 22 | 137 ± 27 | 0.80 | 0.95 | 6 | 129 ± 21 | 1.11 | 0.84 | 16 |
| 6 | 135 ± 19 | 1.10 | 0.64 | 22 | 125 ± 13 | 1.74 | 0.67 | 6 | 139 ± 20 | 0.99 | 0.63 | 16 |
| 7 | 143 ± 13 | 1.07 | 0.63 | 36 | 137 ± 14 | 1.25 | 0.78 | 13 | 146 ± 11 | 1.14 | 0.61 | 21 |
| 8 | 145 ± 15 | 0.85 | 0.80 | 31 | 145 ± 15 | 0.90 | 0.85 | 10 | 145 ± 15 | 0.88 | 0.68 | 23 |
| 9 | 149 ± 18 | 0.85 | 0.74 | 39 | 150 ± 13 | 0.58 | 0.87 | 10 | 148 ± 20 | 0.89 | 0.74 | 29 |
| 10 | 149 ± 16 | 1.06 | 0.71 | 41 | 143 ± 16 | 0.96 | 0.63 | 12 | 152 ± 15 | 1.14 | 0.74 | 29 |
| 11 | 148 ± 16 | 0.89 | 0.66 | 39 | 144 ± 22 | 0.82 | 0.82 | 7 | 149 ± 15 | 0.89 | 0.62 | 32 |
| 12 | 146 ± 14 | 1.09 | 0.75 | 40 | 143 ± 11 | 1.56 | 0.92 | 8 | 146 ± 14 | 1.04 | 0.72 | 33 |
| 13 | 149 ± 15 | 0.74 | 0.64 | 39 | 148 ± 16 | 0.88 | 0.81 | 17 | 152 ± 14 | 0.62 | 0.74 | 18 |
| 14 | 145 ± 17 | 0.84 | 0.60 | 48 | 142 ± 19 | 1.00 | 0.69 | 15 | 147 ± 15 | 0.84 | 0.69 | 29 |
| 15 | 151 ± 13 | 0.75 | 0.76 | 42 | 156 ± 15 | 0.63 | 0.74 | 9 | 150 ± 13 | 0.80 | 0.75 | 34 |
| 16 | 150 ± 14 | 0.79 | 0.69 | 47 | 146 ± 13 | 0.75 | 0.66 | 16 | 152 ± 14 | 0.86 | 0.70 | 32 |
| 17 | 146 ± 14 | 0.95 | 0.76 | 40 | 146 ± 15 | 0.86 | 0.71 | 15 | 146 ± 13 | 0.96 | 0.76 | 26 |
| 18 | 152 ± 13 | 1.05 | 0.62 | 43 | 150 ± 15 | 1.37 | 0.68 | 10 | 152 ± 13 | 0.91 | 0.65 | 33 |
| 19 | 154 ± 13 | 0.80 | 0.61 | 43 | 152 ± 7 | 1.18 | interp | - | 155 ± 14 | 0.81 | 0.63 | 38 |
| 20 | 150 ± 13 | 1.00 | 0.61 | 42 | 152 ± 16 | 1.01 | 0.93 | 7 | 149 ± 13 | 0.93 | 0.69 | 31 |
| 21 | 153 ± 12 | 0.85 | 0.61 | 50 | 158 ± 11 | 1.08 | 0.75 | 17 | 150 ± 12 | 0.70 | 0.67 | 29 |
| 22 | 156 ± 13 | 0.83 | 0.65 | 58 | 154 ± 16 | 0.44 | 0.83 | 6 | 156 ± 13 | 0.84 | 0.70 | 50 |
| 23 | 157 ± 16 | 0.85 | 0.76 | 47 | 157 ± 17 | 0.59 | 0.63 | 12 | 157 ± 16 | 0.95 | 0.82 | 35 |
| 24 | 153 ± 13 | 0.65 | 0.65 | 36 | 152 ± 12 | 0.66 | 0.71 | 8 | 153 ± 14 | 0.62 | 0.59 | 29 |
| 25 | 153 ± 16 | 0.79 | 0.67 | 41 | 150 ± 14 | 1.03 | 0.79 | 10 | 155 ± 16 | 0.72 | 0.64 | 31 |
| 26 | 150 ± 16 | 0.90 | 0.70 | 40 | 158 ± 13 | 0.67 | 0.98 | 6 | 150 ± 16 | 0.96 | 0.76 | 31 |
| 27 | 149 ± 17 | 0.80 | 0.77 | 38 | 145 ± 16 | 1.12 | 0.86 | 10 | 151 ± 17 | 0.68 | 0.72 | 29 |
| 28 | 150 ± 18 | 1.05 | 0.76 | 45 | 153 ± 23 | 0.84 | 0.98 | 7 | 150 ± 17 | 1.11 | 0.75 | 38 |
| 29 | 150 ± 15 | 0.86 | 0.61 | 49 | 143 ± 16 | 0.73 | 0.83 | 13 | 152 ± 13 | 1.00 | 0.62 | 36 |
| 30 | 152 ± 16 | 0.95 | 0.70 | 52 | 151 ± 16 | 0.65 | 0.80 | 15 | 152 ± 17 | 1.08 | 0.75 | 34 |

For older patients (>30 days), we developed regression models to predict the appropriate QTcAd formula parameters using a custom Matlab script. First, age specific 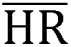 and |m| values were estimated from bins with widths expanded day-by-day until at least n=30 ECGs were included. Within each bin, outlier ECGs were iteratively removed until the HR-QT interval correlation coefficient reached R^2^ > 0.6. Sex-and race-specific differences in HR and the QT interval were tested, but the impact on the regression model was negligible (**Figure 2B**).

Therefore, for older patients, both |m| and 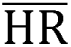 were modeled as a sole function of age and expressed as:

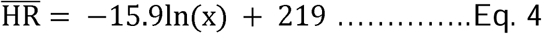

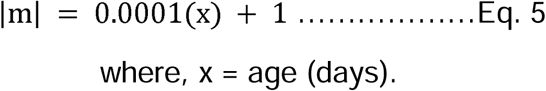

### Age-Based Dynamic QTcAd Thresholding for Pediatric Congenital LQTS Screening

Age-specific QTcAd reference limits were derived from the CNH CVD-free cohort using age-binned estimates of the mean and standard deviation across development. Dynamic lower and upper reference limits were constructed to define normal QTcAd values. A prolonged QTcAd screening threshold was established by adding an empirically optimized margin above the age-specific upper reference limit. Values between the lower reference limit and prolonged threshold were classified as borderline. The optimal margin was selected in the derivation subset of clinically confirmed congenital LQTS patients and the CNH CVD-free control cohort to maximize diagnostic discrimination while minimizing false-positive classifications. When multiple ECGs were available, classification required threshold fulfillment on at least two ECGs obtained ≥30 days apart. Additional details regarding reference limit construction, parameter optimization, borderline/prolonged definitions, and classification rules are provided in the Supplement (**Supplemental Material, Figure S1, Table S1-S2**).

### Calibration of QTcAd Thresholds for Emergency Department ECG Screening

ECGs obtained in the ED may reflect transient physiological stress (e.g., tachycardia, fever, or pain), as such, a separate operating point was calibrated for this single-ECG screening context. Threshold parameters were optimized using the PHN healthy cohort to achieve a false-positive rate and prolonged QT classification frequency comparable to standard Bazett-based screening thresholds. The resulting age-adjusted thresholds were then applied to the CNH ED cohort to compare borderline and prolonged classifications that would prompt repeat ECG testing in clinical practice. Detailed calibration procedures, parameter searches, and final threshold values are provided in the **Supplemental Material**.

### Congenital LQTS screening calculator

To calculate the enhanced QTcAd value from heart rate, QT interval, age (in days), race (white, black, other), and sex (male, female) – we developed a calculator that is optimized for individuals 1 day–23 years old (https://adaptive-qtc-calculator.streamlit.app/). The calculator classifies patients for congenital LQTS screening by implementing the aforementioned optimal threshold algorithm. The publicly accessible code is available (https://github.com/kazihaq/Adaptive-QTc-Calculator).

## Results

### Screening for QTc prolongation in the CNH CVD-free (control) cohort

In CNH CVD-free cohort, the majority of patients were classified as normal by both QTcB and QTcAd formulae **(Figure 3A).** In the all ECG group, QTcB classified 89% of cases as normal and 1.1% as prolonged QTc, and QTcAd classified 81.7% of cases as normal and 3.1% as prolonged QTc. Cross-classification based on a confusion matrix demonstrated agreement between the two formulae, with 88.7% of cases classified as normal by both QTcB and QTcAd **(Supplemental Table S3)**. In the single-ECG subgroup, prolonged QTc classifications were minimal for both QTcAd (0.6%) and QTcB (0.2%) – but, increased slightly in the multiple ECG subgroup (8.4% QTcAd, 2.7% QTcB; **Figure 3A**).

**Figure 3.**
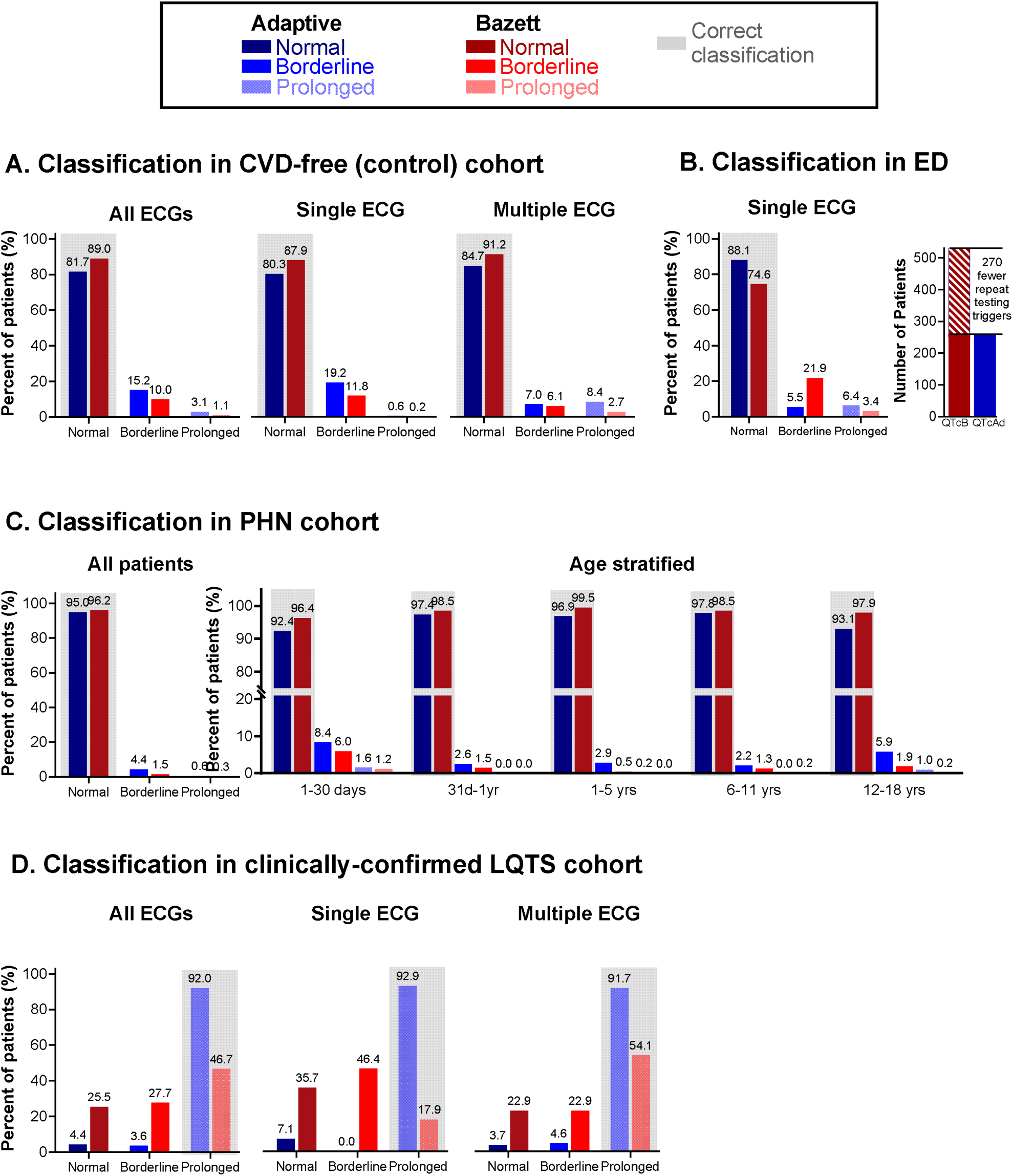
Performance evaluation of adaptive QTc-based congenital LQTS screening. **A)** Percentage of CNH CVD-free (control) subjects (n=4,556) classified as normal, borderline, or prolonged by QTcB and QTcAd in the overall cohort, single-ECG subgroup (n=3,064) and multi-ECG subgroup (n=1,492). **B)** Percentage of ED patients (n=2,002) classified as normal, borderline, or prolonged using QTcB and QTcAd. Inset shows the total number of repeat-testing triggers (borderline +prolonged). **C)** Percentage of PHN healthy children classified as normal, borderline, or prolonged using QTcB and QTcAd. The left panel shows overall classification; the right panel shows age-stratified results (1–30 days (n=251), 31 days–1 year (n=271), 1–5 years (n=421), 6–11 years (n=543), and 12–18 (n=908) years). **D)** Percentage of patients classified as normal, borderline, or prolonged by QTcB and QTcAd in the overall congenital LQTS validation cohort (n=137), the single-ECG subgroup (n=28), and the multi-ECG subgroup (n=109). Note: The correct classification is highlighted in gray (e.g., “normal” for CVD-free patients). Percentage of patients are shown above the bars.

### Screening for QTc prolongation in CNH ED cohort

In the CNH ED cohort, QTcB classified 74.6% of cases as normal (n=1,494), 21.9% as borderline (n=439), and 3.4% as prolonged QTc (n=69), corresponding to an overall positive screen rate (borderline or prolonged) or 25.4%. Using the ED-derived operating point (Δ_ED_ = 23 ms; LowerShift__ED_=14 ms), QTcAd classified 88.1% of cases as normal (n=1,764), 5.5% as borderline (n=110), and 6.4% as prolonged QTc (n=128), reducing the positive screen rate to 11.9%. Overall, QTcAd yielded 270 fewer cases classified as borderline or prolonged (**Figure 3B)**. Cross-classification analysis using a confusion matrix showed that 78.6% of QTcB borderline cases and 20.3% of prolonged cases were classified to normal by QTcAd. Conversely, among QTcAd classifications, only 1.9% of borderline and 4.0% of prolonged cases were classified as normal using QTcB **(Supplemental Table S4)**.

### Screening for QTc prolongation in Pediatric Heart Network cohort

Cross-classification demonstrated high overall agreement in this healthy single-ECG cohort, with 96.1% of cases classified as normal by both QTcB and QTcAd **(Supplemental Table S5)**. Specifically, QTcB classified 98.4% of cases as normal, 1.4% as borderline, and 0.3% as prolonged QTc **(Figure 3C)**. QTcAd classified 95.0% of cases as normal, with a modest increase in borderline (4.4%) and prolonged (0.6%) classifications. Age-stratification showed that neonates were most frequently classified as borderline (6% QTcB, 8.4% QTcAd) or prolonged (1.2% QTcB, 1.6% QTcAd). Other age groups had lower proportions of borderline (0.5-1.9% QTcB, 2.2-5.9% QTcAd) and prolonged classifications (0-1.2% QTcB, 0-1.6% QTcAd). Using prolonged QTc as a positive threshold in this non-event cohort, specificity was 99.7% for QTcB (2,388/2,394 cases, 0.3% false positive) and 99.4% for QTcAd (2,380/2,394, 0.6% false positive).

### Performance evaluation of QTcAd based congenital LQTS screening

The performance of QTcAd and QTcB was evaluated in a testing cohort of 137 confirmed LQTS patients. Using adaptive age-based thresholds, a larger proportion of prolonged QTc cases were correctly classified using the QTcAd (92%, 126/137) compared with QTcB (46.7%, 64/137; **Figure 3D)**. QTcB incorrectly classified 25.5% of cases as normal and 27.7% as borderline, whereas QTcAd incorrectly classified only 4.4% of cases as normal and 3.6% as borderline. Cross-classification showed that 77.1% of QTcB normal cases and 92.1% of borderline cases were correctly reclassified as prolonged by QTcAd **(Supplemental Table S6).** Among patients with only a single ECG (n=28), QTcAd identified 92.9% as prolonged QTc, compared with 17.9% using QTcB **(Figure 3D)**. Overall, QTcAd substantially reduced false-negative classifications in the LQTS cohort. In the full ECG dataset, the false-negative rate decreased from 53.2% under QTcB (25.5% normal + 27.7% borderline) to 8% under QTcAd (4.4% normal + 3.6% borderline).

Lead-time analysis identified a temporal difference in LQTS detection between QTcAd and QTcB among patients classified as prolonged QTc. QTcAd identified prolonged QTc earlier than QTcB in 43.8% of patients, whereas QTcB identified cases earlier in only 1.6%; the remainder were identified on the same day **(Figure 4A).** Paired lead-time comparisons (n=64) demonstrated a longer diagnostic lead time with QTcAd compared with QTcB **(Figure 4B)**. Overall diagnostic performance is provided in **Supplemental Table S7**.

**Figure 4.**
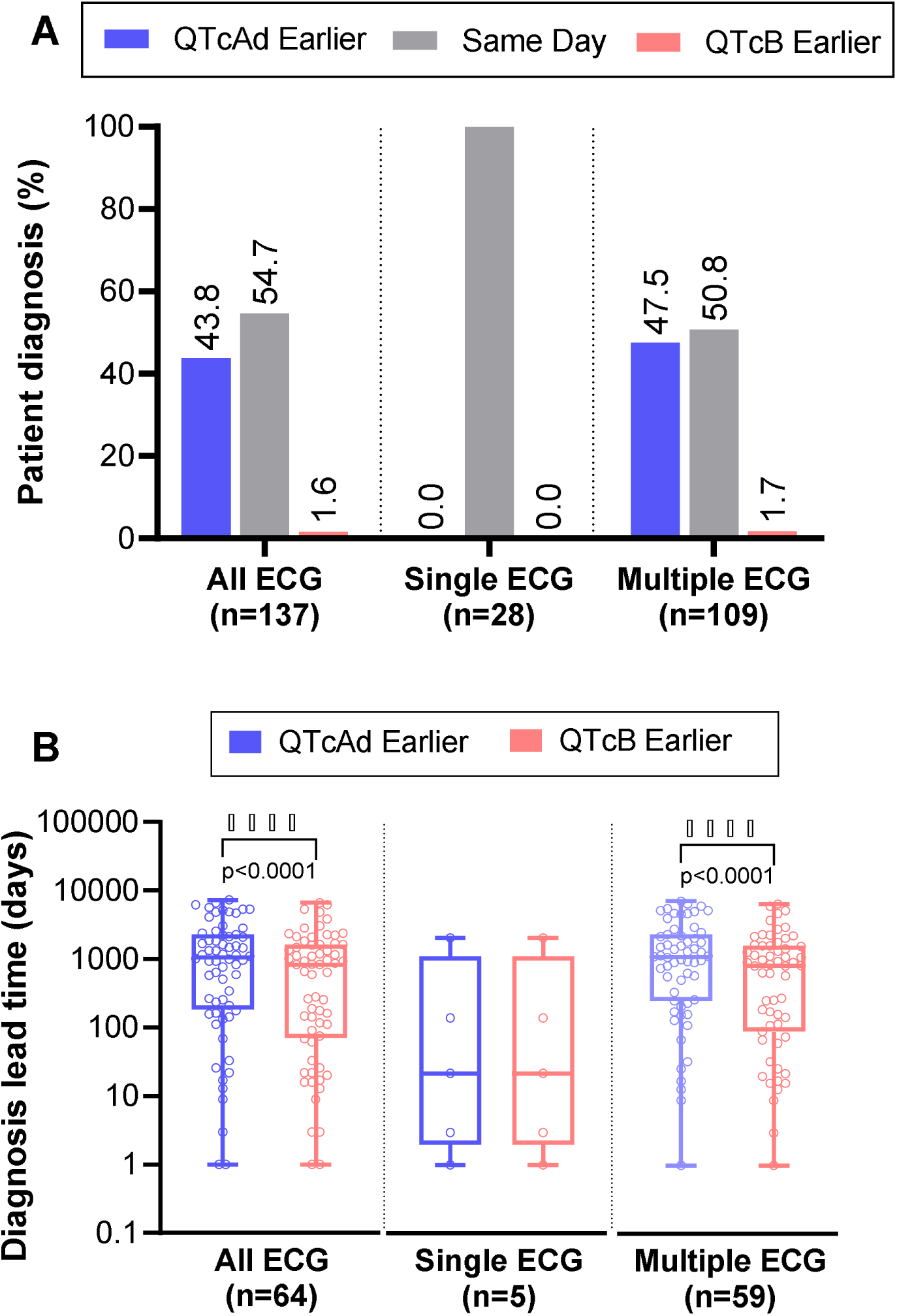
Lead-time difference in patient LQTS diagnosis. **A)** Patients classified with prolonged QT using both Bazett (QTcB) and Adaptive (QTcAd) correction formulae. Bar plots show the percentage of patients diagnosed at an earlier timepoint when using the QTcAd or QTcB correction formulae (or no difference, same day). **B)** Box-and-whisker plots show individual patient lead-time to diagnosis (days from ECG to clinical LQTS confirmation) using QTcB vs QTcAd correction formulae. Datasets include all ECG (n=64), single ECG (n=5), and multiple ECG (n=59) – included data was restricted to patients that were classified as “prolonged” using both methods. Statistical comparison was performed using paired t-test (Wilcoxon signed-rank test); ****p<0.0001

Using the CVD-free control cohort as the non-event population and the confirmed LQTS cohort as the event population, QTcAd demonstrated higher sensitivity (92.0%) than QTcB (46.7%), while maintaining high specificity (96.9% and 98.9%, respectively). Accordingly, QTcAd achieved a higher Youden index (0.889 vs 0.456), indicating improved overall diagnostic discrimination.

## Discussion

In this study, we developed and validated an adaptive QT correction approach (QTcAd) with age-adjusted dynamic thresholds for pediatric congenital LQTS screening. Across multiple independent cohorts, QTcAd improved detection of clinically confirmed LQTS compared with Bazett correction (QTcB), with a marked increase in sensitivity (92.0% vs 46.7%) while maintaining high specificity. This improvement was driven by a reduction in false-negative classifications and earlier detection of prolonged QTc. In parallel, QTcAd reduced the burden of borderline/prolonged classifications in real-world screening settings, yielding fewer repeat-testing triggers in the emergency department cohort. Collectively, these findings support the use of adaptive, age-dependent QTc correction and thresholding to improve diagnostic accuracy and clinical efficiency in pediatric QTc assessment.

### Demographically enhanced QTcAd formula for pediatric populations

Consistent with prior work (Haq et al., 2025), age was the dominant determinant of HR and QT interval in this cohort, with minimal independent contributions from sex, race, or socioeconomic status outside of specific developmental windows. Race interaction affected |m| and 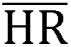 only in the neonates, and therefore day-specific parameter values were derived for this age group. For older patients (infants, children, young adults) the regression models effectively estimate |m| and 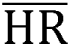 for any given age (in days) until 23 years old. Incorporation of additional demographic variables produced minimal deviation in model trajectories, indicating that age alone sufficiently captures the primary physiological variation. Based on these findings, the empirical model includes age as the sole predictive variable. The current study also expands upon our prior work by improving temporal resolution and extending applicability through early adulthood.

Prior studies have demonstrated age-dependent variation in ECG parameters across both human and experimental models (Haq et al., 2023, 2024; Lue et al., 2022; Schwartz et al., 2002; Swift et al., 2020). Additional demographic factors, such as sex, race, and socioeconomic status, have been associated with ECG variation in healthy populations (Carnevali et al., 2023; Davis et al., 2022; Kaur et al., 2024; Maksimov et al., 2023; Santhanakrishnan et al., 2016; SIMONSON et al., 1960; Taneja et al., 2001). In the CNH CVD-free cohort, race effects were limited to neonates, while sex-related differences emerged primarily during late childhood and adolescence, consistent with prior reports (Bratincsák et al., 2020; Saarel et al., 2018). The childhood opportunity index demonstrated minimal association with ECG parameters, consistent with previous findings (Kézdi et al., 2025).

In prior work (Haq et al., 2025), QTcAd outperformed traditional QT correction formulae by eliminating the inverse HR-QT relationship. In the present study, we extend these findings by demonstrating consistent performance across diverse pediatric populations, including CVD-free individuals, emergency department patients, and confirmed LQTS cases. QTcAd therefore represents a demographically adaptive QT correction approach designed for pediatric populations. Of importance, age-selective comparisons demonstrated only a modest association between COI and the ECG parameters; these effects were small and did not significantly alter the age-dependent trajectories of heart rate or the HR-QT regression slope that form the basis of QTcAd. Thesefindings suggest that developmental physiology remains the dominant determinant of QT adaptation across childhood.

### Congenital LQTS Screening in Pediatric Patients

Congenital LQTS poses a significant health risk in pediatric populations (Balestra et al., 2024; Schwartz et al., 2009), and QTc remains a central biomarker for screening. However, optimal QTc thresholds remain variable across studies and guideline recommendations ((Ackerman et al., 2011; Johnson & Ackerman, 2009; Mazzanti et al., 2018; Mönnig et al., 2006, 2006; Nosetti et al., 2024; Priori et al., 2013, 2015; Schwartz et al., 2009; Vink et al., 2018, 2018; Zeppenfeld et al., 2022), **Table 4**). This variability reflects differences in population demographics, QT measurement, and correction formulae applied (Kanniainen et al., 2024; Vink et al., 2017, 2018).

**Table 4:**
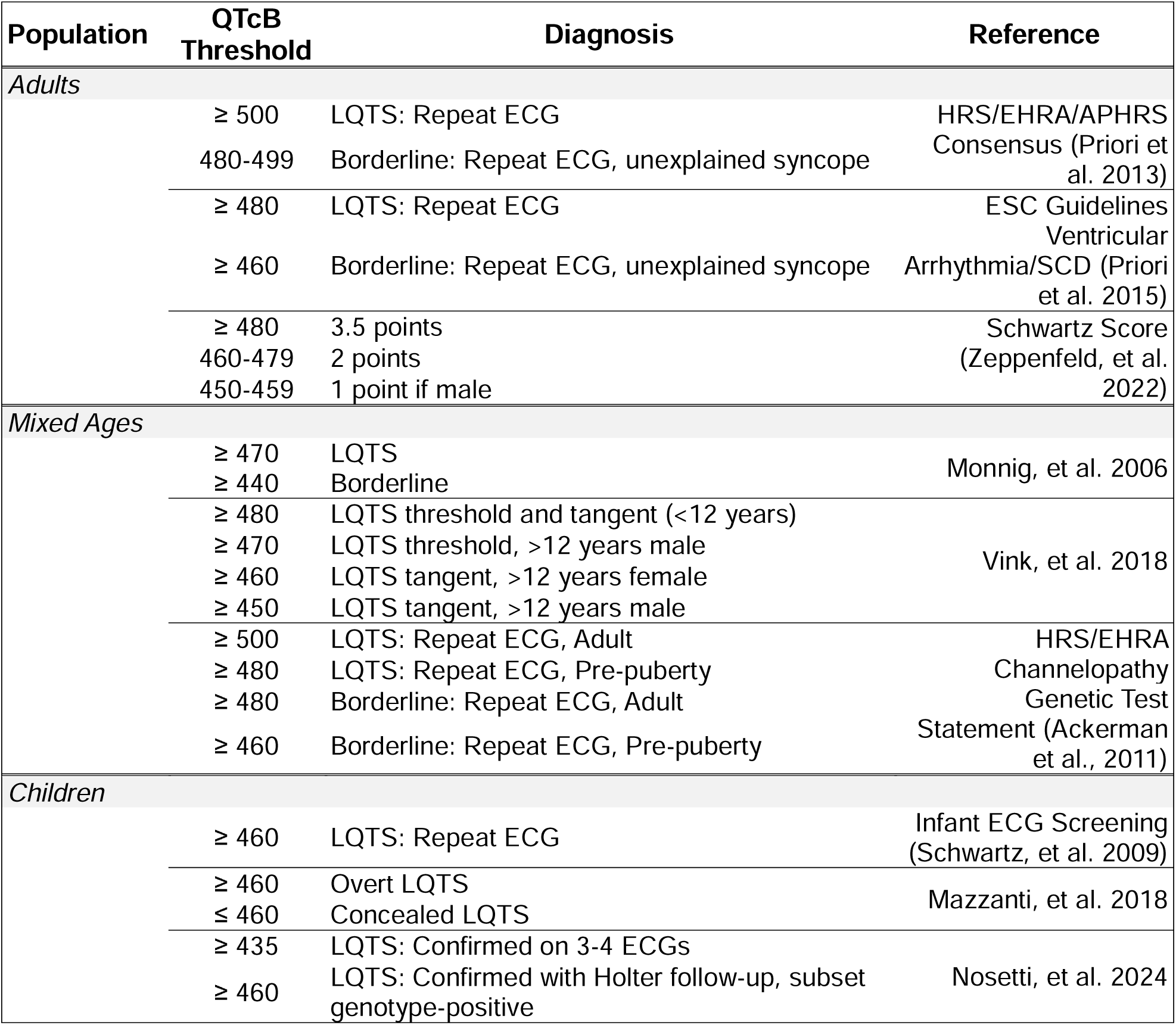
Previously reported QTc cutoff values using Bazett QT correction.

Unlike fixed-threshold approaches, the QTcAd framework applies an age-adjusted dynamic threshold that adapts continuously across development, improving classification accuracy. The limitations of fixed QTc thresholds have been previously noted (Hnatkova et al., 2019; Johnson & Ackerman, 2009; Leppänen et al., 2025; Vink et al., 2018). For example, a neonatal screen study using 460 ms QTcB cutoff reported a positive predictive value of only 39.5% (Stramba-Badiale et al., 2018), while studies in adults have demonstrated high false-positive rates with lower fixed thresholds (Chan et al., 2007). These findings underscore the challenge of distinguishing physiological from pathological QT prolongation when using static thresholds.

Diagnostic uncertainty remains a major challenge in LQTS, with reported misdiagnosis rates and limitations in genetic testing sensitivity (Asatryan et al., 2024; Bains et al., 2023). Many patients referred for evaluation based on QTc values do not harbor pathogenic variants, suggesting that current thresholds may lack specificity (Brogger et al., 2024). An adaptive QTc framework may help address these limitations by aligning thresholds with developmental physiology. Notably, the CNH cohort was intentionally defined using clinically confirmed congenital LQTS rather than restricting inclusion to genotype-positive cases. This approach reflects real-world practice, where a substantial subset of patients demonstrate strong phenotypic evidence of LQTS despite negative or inconclusive genetic testing. For convenience, an automated calculator is available to facilitate rapid, standardized implementation of this approach in clinical and research settings (https://adaptive-qtc-calculator.streamlit.app/).

### Age-adjusted dynamic QTcAd threshold outperforms fixed QTcB threshold for congenital LQTS screening

QTcAd-based age-adjusted dynamic thresholding demonstrated improved performance for pediatric congenital LQTS screening compared with fixed QTcB thresholds. Using LQTS patients as the event population and the CVD-free cohort as the non-event population, QTcAd improved sensitivity while maintaining high specificity, resulting in a higher Youden index. This improvement was primarily driven by a reduction in false-negative classifications, with the proportion of LQTS patients classified as normal or borderline decreasing from 53.2% with QTcB to 8.0% with QTcAd. Importantly, this improvement did not result in a substantial increase in false-positive classifications in control populations, where most individuals remained classified as normal. In a real-world screening setting, application of QTcAd thresholds in the ED cohort reduced borderline/prolonged classifications that would require follow-up, resulting in 270 fewer repeat-testing triggers. These findings highlight the limitations of fixed QTc thresholds and support the use of age-adaptive thresholding to improve both diagnostic accuracy and downstream clinical efficacy.

Prior studies evaluating QTcB performance in pediatric populations have reported variable results, with some demonstrating increased false-positive classifications and others showing comparable performance relative to alternative correction methods (Andršová et al., 2020; Dahlberg et al., 2021; Leppänen et al., 2025; Stramba-Badiale et al., 2018; Van Dorn et al., 2011). A consistent observation across studies is the instability of QTcB classifications over time, with initially prolonged values often normalizing on follow-up evaluation (Leppänen et al., 2025; Van Dorn et al., 2011). These limitations likely reflect the lack of demographic adjustment and reliance on fixed thresholds within conventional QTcB-based screening approaches. Alternative pediatric-specific QT correction formulae have also been proposed (Benatar & Feenstra, 2015; Phan et al., 2015; Wernicke et al., 2005), although some may overestimate QTc values (Haq et al., 2025). In contrast, QTcAd incorporates demographic adaptability and dynamic thresholding, providing a more physiologically aligned framework for pediatric QTc assessment. Importantly, this study was designed to evaluate QTcAd relative to clinically implemented QTcB screening workflows that rely on fixed thresholds, rather than to isolate the independent effects of thresholding versus correction methodology.

## Limitations

Although we evaluated the effects of sex, race, and childhood opportunity index (COI) on ECG parameters, the impact of these variables on the final QTcAd model was limited, and additional demographic and environmental factors not captured in this study may contribute to variability in the HR-QT relationship. Race categorization was broad, and COI data were derived primarily from the Washington, DC–Maryland–Virginia region, which may limit generalizability. In this cohort, the influence of COI on ECG parameters was modest and age-specific; however, socioeconomic factors may affect other cardiovascular biomarkers and warrant further investigation in larger and more geographically diverse populations. The study population was restricted to pediatric and young adult subjects, and additional work is needed to determine the applicability of QTcAd in older adult populations. The congenital LQTS cohort consisted of clinically confirmed cases rather than exclusively genotype-positive patients, reflecting contemporary diagnostic practice but potentially introducing greater phenotypic heterogeneity. Finally, QTcAd threshold performance was evaluated primarily using data from a single center, and multicenter validation will be important to confirm generalizability.

## Conclusion

This study extends a previously developed age-based QT correction by incorporating additional demographic parameters and expanding applicability across the pediatric age spectrum. An age-adjusted, QTcAd-based dynamic threshold demonstrated improved detection of congenital LQTS compared with fixed QTcB thresholds, while reducing excess QT prolongation classifications in non-event populations. These findings support the adoption of adaptive, demographically informed QTc correction and thresholding to improve diagnostic accuracy and reduce unnecessary downstream evaluation in pediatric QTc screening.

## Data Availability

All data produced in the present work are contained in the manuscript

## Acknowledgements

Acknowledgements:We thank Jennifer Klein from the Division of Cardiology for valuable scientific insight regarding childhood opportunity index classifications, and Grace Zereski for assisting with manual validation of clinically confirmed LQTS cases identified with DataBricks. We also acknowledge the NIH/NHLBI Pediatric Heart Network Echo-Z/Normal ECG dataset, which was used in preparation of this work. Data was downloaded from www.pediatricheartnetwork.org on 02/11/2025.

## Funding

This work was supported by the National Institutes of Health grant R01HD108839 (NGP), Children’s National Heart & Lung Center, The Sheikh Zayed Institute for Pediatric Surgical Innovation, and the Foglia, Hills, and Seelig families.

## Supplemental Materials

Supplemental Methods, Supplemental Figure S1, Supplemental Tables S1-7

## Supplemental Material

### Supplemental Methods

#### Age-Based Dynamic QTcAd Thresholding for Pediatric Congenital LQTS Screening

We aimed to establish a dynamic, age-adjusted threshold to screen pediatric patients for congenital LQTS. Age-specific reference limits were derived from the CNH CVD-free control cohort by estimating the mean (μ) and standard deviation (σ) of QTcAd within age bins (126 age bins spanning 1–8,300 days). The upper reference limit was defined as Upper(age) = μ(age) + 1.645·σ(age), corresponding to the 95th percentile under a normal approximation, and the lower reference limit as Lower(age) = μ(age) − 1.645·σ(age) **(Supplemental Figure S1).** To define the screening threshold for prolonged QTcAd, a fixed margin Δ was added above the upper reference limit to indicate prolonged repolarization when QTcAd ≥ Upper(age) + Δ. Values between the lower reference limit and this prolonged threshold (Lower(age) ≤ QTcAd < Upper(age) + Δ) were classified as borderline, and values below the lower limit were classified as normal.

The optimal margin Δ = 15 ms was determined empirically using the derivation subset of confirmed LQTS patients and the CNH CVD-free control cohort. Δ values from 5–25 ms were evaluated by examining the trade-off between prolonged classification rates in the LQTS derivation cohort (proxy for sensitivity) and the control cohort (proxy for false-positive rate) **(Supplemental Figure S1)**. When multiple ECGs were available, classification required the criterion to be met on ≥ 2 ECGs separated by ≥ 30 days. If only a single ECG was available, classification was based on the single ECG. The complete rule-based classification protocol and schematic threshold definitions are provided **(Supplemental Tables S1-S2).**

#### Calibration of QTcAd Thresholds for Emergency Department ECG Screening

For the ED cohort, a single-ECG, patient-level classification protocol was applied. QTcB (Bazett) thresholds follow standard definitions: prolonged if QTcB ≥470 ms (extreme flag if ≥490 ms), borderline if 440-469 ms, and normal if <440 ms. Both borderline and prolonged classifications were considered repeat-testing triggers, reflecting real-world clinical practice in which finding either may prompt repeat ECG acquisition for further evaluation. However, ECGs obtained in the ED represent a clinically biased population, since they are often recorded under acute physiological stress (e.g., tachycardia, fever, or pain). Applying the disease-screening thresholds derived for stable populations (see prior section) in this context could inflate repeat-testing Accordingly, an ED-specific operating point was defined to better reflect this use case. To accomplish this task, parameter calibration was performed using the PHN healthy single-ECG cohort as a reference population to estimate the false-positive burden under normal conditions. In this cohort, QTcB yielded a false-positive rate (borderline + prolonged) of 1.75%, including 0.25% classified as prolonged. A grid search was conducted over candidate parameter values (Δ_ED_: 1-60 ms; LowerShift__ED_: 1-40 ms) to identify combinations for QTcAd that would 1) maintain a false-positive rate comparable to QTcB, and 2) maintain a comparable prolonged classification rate.

The optimal ED operating point was Δ_ED_ = 23 ms and LowerShift__ED_ = 14 ms. Under these parameters, QTcAd achieved a false-positive rate of 1.67% in the PHN cohort (borderline 1.42%, prolonged 0.25%; normal 98.33%), thus satisfying both calibration criteria. As such, QTcAd classification in the ED cohort was defined as: Prolonged: QTcAd ≥ Upper(age) + 23 ms; Borderline: Lower(age) + 14 ms ≤ QTcAd < Upper(age) + 23 ms and, Normal: QTcAd < Lower(age) + 14 ms. The same repeat-testing trigger (borderline or prolonged) was applied to both QTcB and QTcAd to enable direct comparison of repeat-ECG burden in the ED cohort. In a real-world ED screening context, the calibrated QTcAd thresholds may reduce unnecessary repeat-ECG triggers while maintaining detection of clinically significant QT prolongation at a rate comparable to standard QTcB criteria.

## Supplemental Figure

**Figure S1.**
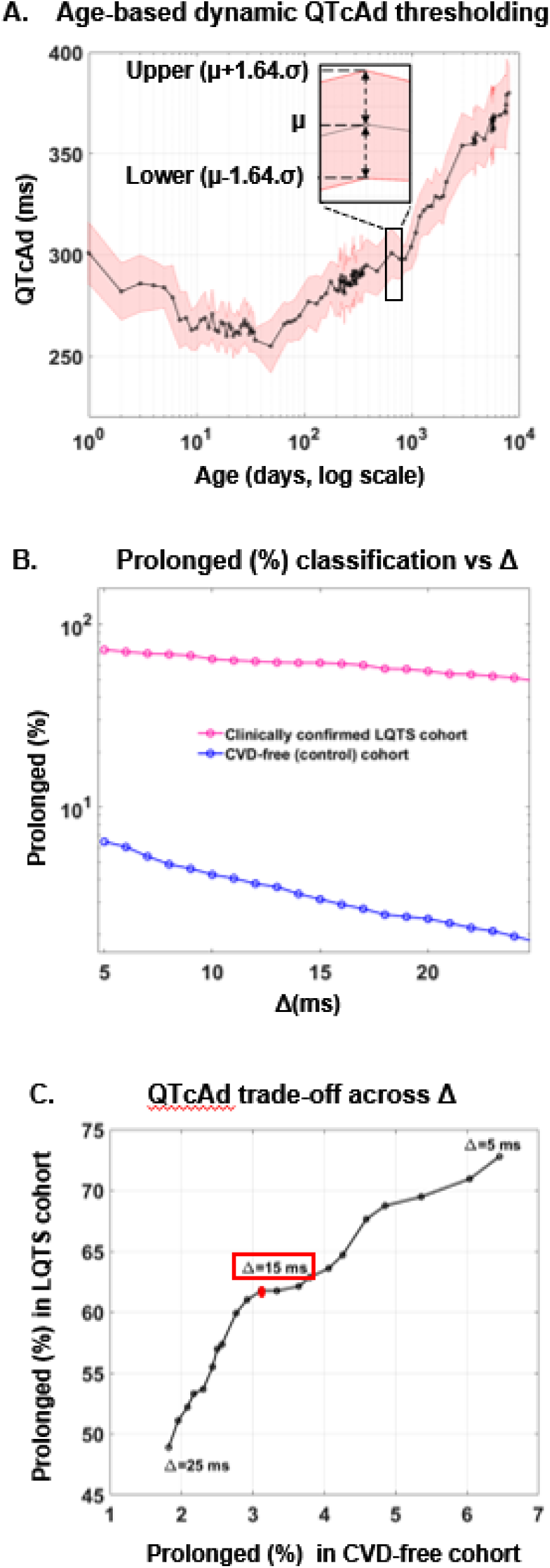
**A)** Age-based dynamic QTcAd thresholding for congenital LQTS screening. QTcAd values from CVD-free patients were grouped into 126 age bins spanning 1-8,300 days. For each bin, the mean QTcAd (μ) and standard deviation (σ) were estimated. The solid black curve shows μ(age) and dashed red bounds represent the age-specific reference limits defined as Lower(age) = μ(age) − 1.645σ(age) and Upper(age) = μ(age) + 1.645σ(age). **B)** Effect of Δ on prolonged classification rates. The percentage of patients classified as prolonged by QTcAd is shown as a function of the adaptive margin Δ for the clinically confirmed LQTS cohort and the CVD-free control cohort. Increasing Δ raises the prolonged threshold (Upper(age) + Δ), resulting in a progressive decrease in prolonged classifications in both cohorts. The LQTS curve reflects the proportion of true positives detected, while the control curve approximates the false-positive rate. **C)** Trade-off between sensitivity and false-positive rate across Δ. Each point represents the relationship between the percentage of patients classified as Prolonged in the LQTS cohort (proxy for sensitivity) and the control cohort (proxy for false-positive rate) for a given Δ. As Δ increases, the false-positive rate decreases but sensitivity also declines. The elbow of the curve occurs near Δ = 15 ms, representing the optimal balance between sensitivity and specificity for congenital LQTS screening.

## Supplemental Tables

**Table S1.** Rule-based protocol used for ECG analysis.

| Analysis component | Definition |
| --- | --- |
| Prior to congenital LQTS diagnosis | Only ECGs acquired before the first electronic health record documented LQTS confirmation data were analyzed (acquisition date < first date of clinical confirmation of congenital LQTS) |
| Control cohort | All qualifying ECGs were included |
| Multiple ECGs available | Classification required the criterion to be met on $\geq 2$ ECGs separated by $\geq 30$ days |
| Single ECG available | Classification based on single ECG |

**Table S2.** QTc classifications based on Bazett (QTcB) or Adaptive (QTcAd) formulae. To evaluate for prolonged classification in patients with multiple ECGs, the “any ECG” criterion was first evaluated (QTc ≥ 490 ms). If satisfied, the patient was classified as prolonged. If not satisfied, the patient required at least 2 ECGs (separated by at least 30 days) to satisfy the criterion. For borderline classification, patients with multiple ECGs were evaluated if they did not meet the prolonged criteria. For patients with only a single ECG recorded, classification was based on that available ECG. QTcB includes fixed threshold. QTcAd includes age-adjusted percentile-based limits. As discussed in the methods section, QTcAd formulat has four components: 1) upper(age) defined as: μ(age) + 1.645 σ(age), 2) lower(age) defined as: μ(age) - 1.645 σ(age), 3) optimized margin “Δ” defined as: 15 ms, and 4) prolonged QTc threshold defined as: upper(age) + Δ

| Formula | Classification | ECGs | Criteria |
| --- | --- | --- | --- |
| *QTcB - Bazett* |  |  |  |
| | Prolonged | Any<br>$\geq 2$<br>1 | $QTcB \geq 490$ ms<br>$\geq 2$ ECGs with $QTcB \geq 470$ ms, separated by $\geq 30$ days<br>$QTcB \geq 470$ ms |
| | Borderline | $\geq 2$<br>1 | $\geq 2$ ECGs with $440 \text{ ms} \leq QTcB \leq 470 \text{ ms}$, separated by $\geq 30$ days<br>$440 \text{ ms} \leq QTcB \leq 470 \text{ ms}$ |
| | Normal | All | $QTcB < 440$ ms |
| *QTcAd - Adaptive* |  |  |  |
| | Prolonged | $\geq 1$ | Any ECG with $QTcAd \geq \text{upper (age)} + \Delta$ |
| | Borderline | $\geq 2$<br>1 | $\geq 2$ ECGs with $\text{lower (age)} \leq QTcAd < \text{upper (age)} + \Delta$, separated by $\geq 30$ days<br>$\text{Lower (age)} \leq QTcAd < \text{upper (age)} + \Delta$ |
| | Normal | All | $QTcAd < \text{lower (age)}$ |

**Table S3.** Confusion Matrix in CVD-free (control) cohort (row-normalized and %). QTcB: Bazett, QTcAd: Adaptive, N: Normal, B: Borderline, P: Prolonged QTc classification. Highlighted values show agreement between the two formulae, as both QTcB and QTcAd correctly classify “normal” patients.

|  |  | All (n=4556) |  |  | Single ECG (n=1492) |  |  | Multiple ECG (n=3064) |  |  |
| --- | --- | --- | --- | --- | --- | --- | --- | --- | --- | --- |
| QTcB | N | **3597**<br>**(88.7%)** | 394<br>(9.7%) | 63<br>(1.6%) | **2365**<br>**(87.8%)** | 327<br>(12.1%) | 2<br>(0.1%) | **1232**<br>**(90.6%)** | 67<br>(4.9%) | 61<br>(4.5%) |
|  | B | 123<br>(27.1) | 296<br>(65.2%) | 35<br>(7.7%) | 94<br>(25.9%) | 259<br>(71.3%) | 10<br>(2.8%) | 29<br>(31.9%) | 37<br>(40.7%) | 35<br>(27.5%) |
|  | P | 2<br>(4.2%) | 2<br>(4.2%) | 44<br>(91.7%) | 0<br>(0%) | 2<br>(28.6%) | 5<br>(71.4%) | 2<br>(4.9%) | 0<br>(0%) | 39<br>(95.1%) |
|  |  | N | B | P | N | B | P | N | B | P |
|  |  | QTcAd |  |  |  |  |  |  |  |  |

**Table S4.** Confusion Matrix in CNH Emergency Department cohort (row-normalized and %). QTcB: Bazett, QTcAd: Adaptive, N: Normal, B: Borderline, P: Prolonged QTc classification. Number of patients correctly classified as “normal” with QTcAd formula, but misclassified as “borderline/prolonged” by QTcB are highlighted.

|  |  |  |  |  |
| --- | --- | --- | --- | --- |
| QTcB | N | 1405<br>(94%) | 29<br>(1.9%) | 60<br>(4%) |
|  | B | **345**<br>**(78.6%)** | 49<br>(11.2%) | 45<br>(10.3%) |
|  | P | **14**<br>**(20.3%)** | 32<br>(46.4%) | 23<br>(33.3%) |
|  |  | N | B | P |
|  |  | QTcAd |  |  |

**Table S5.** Confusion Matrix in Pediatric Heart Network cohort (row-normalized and %). QTcB: Bazett, QTcAd: Adaptive, N: Normal, B: Borderline, P: Prolonged QTc classification. Number of patients correctly classified as “normal” with QTcAd formula, but misclassified as “borderline” by QTcB are highlighted.

|  |  |  |  |  |
| --- | --- | --- | --- | --- |
| QTcB | N | 2263<br>(96.1%) | 85<br>(3.6%) | 7<br>(0.3%) |
|  | B | **11**<br>**(33.3%)** | 19<br>(57.6%) | 3<br>(9.1%) |
|  | P | 0<br>(0%) | 2<br>(33.3%) | 4<br>(66.7%) |
|  |  | N | B | P |
|  |  | QTcAd |  |  |

**Table S6.** Confusion Matrix in clinically confirmed congenital LQTS cohort (row-normalized and %). QTcB: Bazett, QTcAd: Adaptive, N: Normal, B: Borderline, P: Prolonged QTc classification. Number of patients correctly classified as “prolonged” with QTcAd formula, but misclassified as “normal” by QTcB are highlighted.

|  |  | All (n=137) |  |  | Single ECG (n=28) |  |  | Multiple ECG (n=109) |  |  |
| --- | --- | --- | --- | --- | --- | --- | --- | --- | --- | --- |
| QTcB | N | 6<br>(17.1%) | 2<br>(5.7%) | **27**<br>**(77.1%)** | 2<br>(20%) | 0<br>(0%) | **8**<br>**(80%)** | 4<br>(16%) | 2<br>(8%) | **19**<br>**(76%)** |
|  | B | 0<br>(0%) | 3<br>(7.9%) | 35<br>(92.1%) | 0<br>(0%) | 0<br>(0%) | 13<br>(100%) | 0<br>(0%) | 3<br>(12%) | 22<br>(88%) |
|  | P | 0<br>(0%) | 0<br>(0%) | 64<br>(100%) | 0<br>(0%) | 0<br>(0%) | 5<br>(100%) | 0<br>(0%) | 0<br>(0%) | 59<br>(100%) |
|  |  | N | B | P | N | B | P | N | B | P |
|  |  | QTcAd |  |  |  |  |  |  |  |  |

**Table S7.** Diagnostic Performance of Bazett (QTcB) and Adaptive (QTcAd) formulae; clinically confirmed congenital LQTS cohort.

| Formula | Sensitivity (%) | Specificity (%) | Youden Index |
| --- | --- | --- | --- |
| QTcB | 46.7 | 98.9 | 0.456 |
| QTcAd | 92.0 | 96.9 | 0.889 |

## Notes

### Competing Interest Statement

The authors have declared no competing interest.

### Funding Statement

This work was supported by the National Institutes of Health grant R01HD108839 (NGP), Childrens National Heart & Lung Center, The Sheikh Zayed Institute for Pediatric Surgical Innovation, and the Foglia, Hills, and Seelig families.

### Author Declarations

This retrospective study was approved the the Children's National Hospital IRB (#12146)

